# Robust radiomic signatures of intervertebral disc degeneration from MRI

**DOI:** 10.1101/2025.04.01.25325008

**Authors:** Terence McSweeney, Aleksei Tiulpin, Narasimharao Kowlagi, Juhani Määttä, Jaro Karppinen, Simo Saarakkala

## Abstract

Low back pain (LBP) is the most common musculoskeletal symptom worldwide and intervertebral disc (IVD) degeneration is an important contributing factor. To improve the quantitative phenotyping of IVD degeneration from T2-weighted MRI and better understand its relationship with LBP, multiple shape and intensity features have been investigated. IVD radiomics have been less studied but could reveal sub-visual imaging characteristics of IVD degeneration. The aim of this study was to identify a robust radiomic signature from deep learning segmentations for IVD degeneration classification. We used data from Northern Finland Birth Cohort 1966 members who underwent lumbar spine T2-weighted MRI scans at age 46-47 (n=1397). We used a deep learning model to segment the lumbar spine IVDs and extracted 737 radiomic features, as well as calculating IVD height index and peak signal intensity difference. Intraclass correlation coefficients across image and mask perturbations were calculated to identify robust features. Sparse partial least squares discriminant analysis was used to train a Pfirrmann grade classification model. The radiomics model had balanced accuracy of 76.7% (73.1-80.3%) and Cohen’s Kappa of 0.70 (0.67-0.74), compared to 66.0% (62.0-69.9%) and 0.55 (0.51-0.59) for an IVD height index and peak signal intensity model. 2D sphericity and interquartile range emerged as radiomics-based features that were robust and highly correlated to Pfirrmann grade (Spearman’s correlation coefficients of −0.72 and −0.77 respectively). Based on our findings these radiomic signatures could serve as alternatives to the conventional indices, representing a significant advance in the automated quantitative phenotyping of IVD degeneration from standard-of-care MRI.

## I. Introduction

**L**OW back pain (LBP) is the number one disease accounting for years lived with disability globally [1] and carries a huge disease burden [2]. Intervertebral disc (IVD) degeneration is considered an important contributing factor to LBP [3]–[6] but is also highly prevalent in asymptomatic populations [7]. The specific role of IVD degeneration in LBP may be obscured by the subjective and coarse nature of qualitative IVD degeneration grading from clinical magnetic resonance imaging (MRI). State-of-the-art deep learning (DL) and radiomics-based approaches offer a means to overcome this by extracting more quantitative, granular, and discriminative information from MRI of the IVD, which could facilitate novel insights into the mechanisms of IVD degeneration and its contribution to LBP.

The current approach to degeneration grading relies on visual evaluation using semi-quantitative grading such as Pfirrmann’s five-point scale, which is based on gross structural changes of the IVD [8] (Figure 1). Pfirrmann grade is determined from the mid-sagittal slice of a T2-weighted MRI where the IVD homogeneity, annulus fibrosis-nucleus pulposis distinction, IVD height, and signal intensity are qualitatively determined. The method is subjective, inherently costly of time and expertise, and has variable intra- and inter-rater reliability [9]–[12]. Deep learning (DL) models have been trained to automatically classify Pfirrmann grade [13] but the lack of interpretability and bias to training data means their utility is also limited [13], [14].

**Fig. 1.**
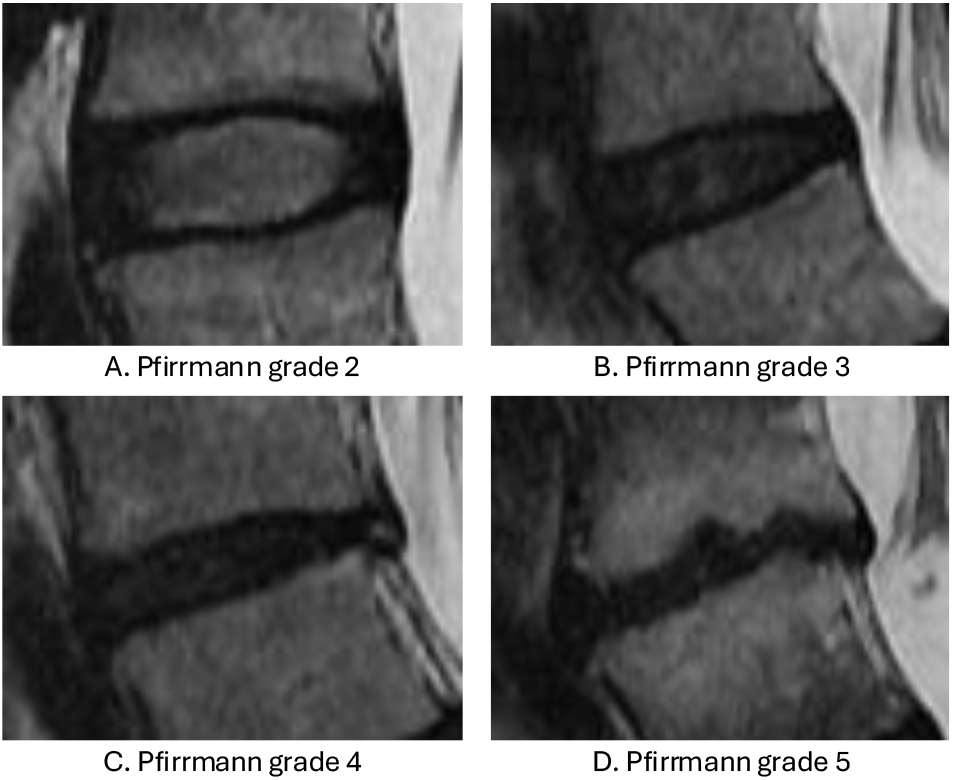
Examples of Pfirrmann grade 2-5 IVD degeneration. **A**. Pfirrmann grade 2 has a hyper-intense but somewhat inhomogeneous nucleus pulposis (NP), clear distinction between the NP and annulus fibrosis (AF), and normal IVD height; **B**. Pfirrmann grade 3 has an inhomogeneous grey NP with intermediate intensity, unclear distinction between the NP and AF, and slight decrease in IVD height; **C**. Pfirrmann grade 4 has a hypo-intense and inhomogeneous grey to black NP, loss of distinction between the NP and AF, and moderate decrease in IVD height; **D**. Pfirrmann grade 5 has a hypo-intense inhomogeneous NP, loss of distinction between the NP and AF, and collapsed IVD space.

Quantitative MRI (qMRI) sequences such as T1*ρ* and T2-maps have been used to detect biochemical and compositional changes in the IVD for better characterisation of degeneration. Features derived from qMRI have shown potential to identify the earliest stages of IVD degeneration [15] but their use is exploratory and they require further validation before widespread implementation [16]. From the most common clinical sequence, T2-weighted MRI, measures of IVD height, e.g. IVD height index [17]–[20], and signal intensity, e.g. peak signal intensity difference [20], [21], are well established features and have been considered candidate imaging biomarkers of IVD degeneration [22]. Such features do not capture the earliest degeneration processes in the IVD, but correspond to key characteristics of the Pfirrmann grades, with loss of nucleus pulposis signal intensity as the first visible sign of degeneration, followed by loss of IVD height [23].

As such, these two conventional indices represent only a fraction of the possible shape, intensity, and texture features that can be extracted from T2-weighted MRI of the IVD. The process of extracting and modelling such large sets of quantitative texture features is referred to as radiomics. The radiomic hypothesis is that a given tissue’s phenotypic, proteomic, and genomic information can be inferred using high throughput analysis of macroscopic imaging features [24]. In the case of the IVD, height and signal intensity capture gross visual characteristics of degeneration but radiomics may reveal more subtle sub-visual changes to improve the objective phenotyping of IVD degeneration and its severity.

Despite an increasing application to musculoskeletal imaging, relatively few studies have investigated radiomics of the IVD from clinical MRI. Most studies quantifying IVD degeneration have tested individual shape or intensity features from T2-weighted MRI, e.g. IVD height [18], [25], fractal dimension [26], T2 signal intensity [27]–[29], and custom shape features [30], although some have used larger sets of texture features [31]–[33]. IVD radiomics from T2-weighted MRI have also been investigated for the classification of painful annular fissures [34], spine surgery prediction [35], and LBP classification [36]–[38].

These studies, whether using conventional indices or radiomics, have relied on manual or semi-automated (non-DL) segmentation of the IVD from which features can be calculated. Additionally, they have small sample sizes and the robustness of the chosen features is under-reported or untested. It is unclear how robust radiomic features are to DL segmentation masks in the context of IVD degeneration classification. Furthermore, it is still unknown whether IVD radiomics can deliver on their purported potential to ‘capture additional information not currently used’ [24] and maximise the value of information in standard of care lumbar spine MRI. This is particularly relevant to clinical sequences such as T2-weighted MRI, which are ubiquitous in spine research and clinical settings, but whose rich data may be underutilised for quantifying IVD degeneration.

The aim of this study is to identify a robust radiomic signature for IVD degeneration using DL segmentation in a population-based cohort. We hypothesize that a subset of radiomic features from T2-weighted IVD MRI can achieve high classification accuracy compared to the conventional indices IVD height index and peak signal intensity difference when using DL segmentation. To test this, we will apply image and mask perturbations to DL segmentations prior to radiomic feature extraction. The radiomic features and conventional indices will then be compared for Pfirrmann grade classification accuracy and reliability. We also aim to examine the features of highest relevance in the resulting model to test for features containing additional information not captured by the conventional indices.

## II. Methods

### A. Data

#### 1) Population

The study population is drawn from the Northern Finland Birth Cohort 1966 (NFBC1966), which is a prospective birth cohort started in 1965. The follow-up data and MRI data used in this study were collected from 2012 to 2014 when participants were between 45 and 47 years of age [39], [40]. Ethics approval for the NFBC1966 was granted by the Northern Ostrobothnia Hospital District Ethics Committee. The population demographics are detailed in Table I and further information regarding the recruitment and population characteristics is available in Nordstrom et al. [39].

**TABLE I.**
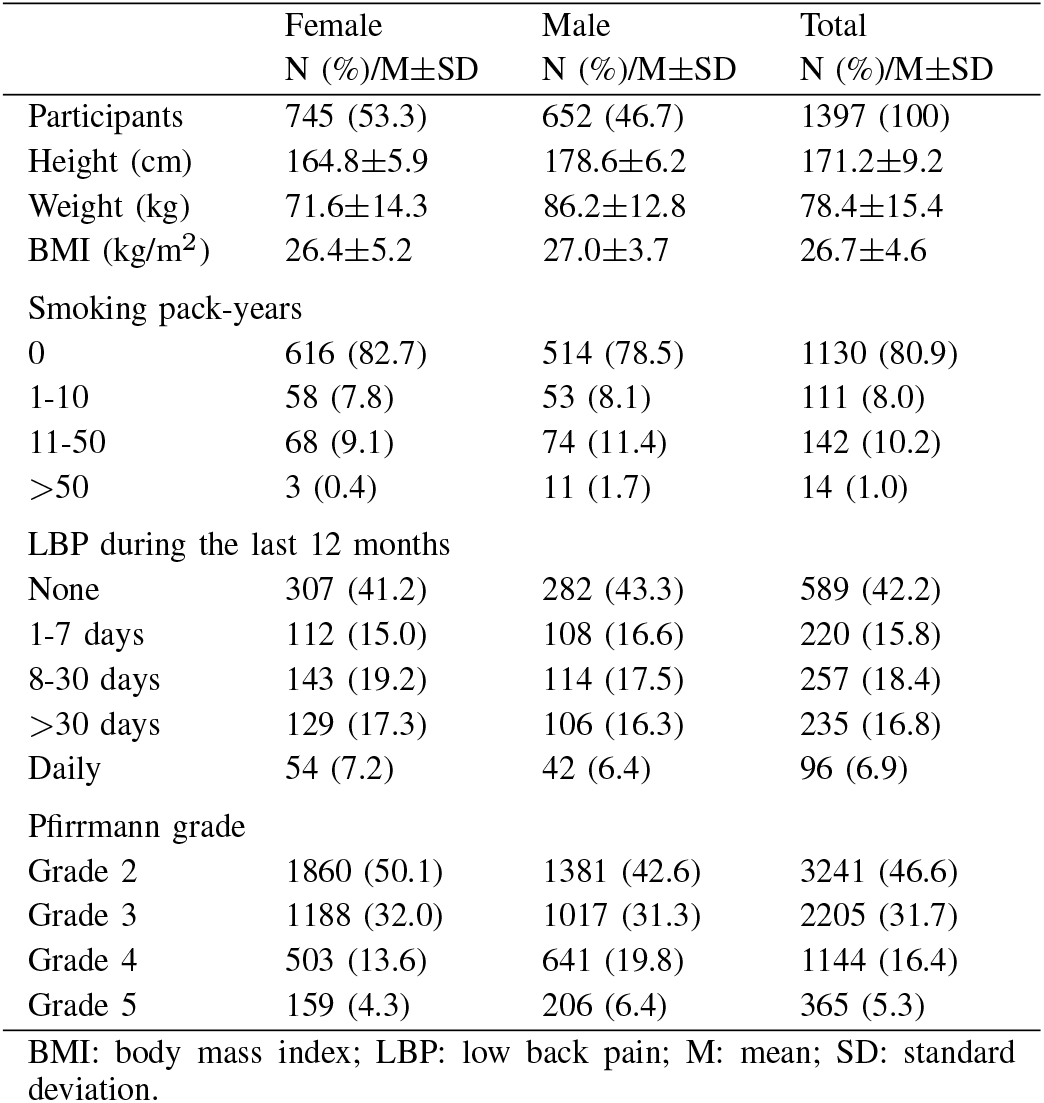
Population characteristics and degeneration prevalence.

#### 2) MRI

The study used T2-weighted sagittal MRI acquired on 1.5-Tesla equipment (Signa HDxt; General Electric, Milwaukee, WI). Sagittal T2-weighted fast-recovery fast spin-echo images were captured with repetition time/effective echo time 3500/112 ms, 4 averages, 280 × 280mm field of view, 512 × 512 slice dimensions, 3mm slice thickness, and 1mm inter-slice gap. Images unsuitable for radiomic feature calculation due to metal artefacts (n=3), developmental anomaly (n=1), severe scoliosis (n=1), and poor image quality or missing sequences (n=24), were removed.

The study sample was limited to those participants for whom consensus IVD degeneration evaluations had been carried out as part of a previous study [9]. For these evaluations, blinded Pfirrmann grading was done by two experienced musculoskeletal radiologists and one physician experienced in spinal imaging (with a combined experience of over 56 years). Pfirrmann grading inter-rater reliability ranged from fair to good (Cohen’s Kappa = 0.39-0.79). A fourth expert rater determined the consensus Pfirrmann grade. Given the extremely low prevalence of Pfirrmann grade 1 at the 46-year follow-up, Pfirrmann grades 1 and 2 have historically been pooled resulting in 4 grades (2-5) available for analysis.

The final dataset consisted of 6985 IVDs from 1397 participants. The prevalence and distribution of degenerative changes is shown in Table I. Data were split into a development set (80%, n=5588 IVDs) and test set (20%, n=1397 IVDs) stratified by participant and Pfirrmann grade.

#### 3) Segmentation

We used an existing DL model trained in the NFBC1966 [41] to segment the lumbar spine IVDs from 4 or 5 sagittal slices centred on the numerical mid-sagittal slice/s. Slices in this range were selected as they captured the middle third of most IVDs and vertebral bodies provided there was not a significant scoliosis. The DL model performance was evaluated against a set of 300 IVDs and their adjoining vertebral bodies from the development set, which were manually annotated by an experienced musculoskeletal researcher (JM). Jaccard index and 95% Hausdorff distance were used to assess the quality of the DL IVD segmentation model. The DL segmentations were manually screened to remove slices where the DL segmentation failed completely.

#### 4) Image-mask perturbations

In preparation for testing the robustness of radiomic features to image-mask variations besides DL segmentation, additional sets of perturbed images and DL masks were generated for the set of 300 manually annotated IVDs in the development set. Random Gaussian noise was applied to the images alone and combined translation and rotation (TR) were applied to the images and masks together. Erosion, dilation, and contour randomisation (CR) were applied to the masks. Combinations of the perturbations were also applied (dilation + TR, erosion + TR, CR + TR, CR + TR + noise) resulting in a final set of 10 perturbations including the DL masks. The settings for the perturbations are detailed in Table II, with examples of the mask perturbations illustrated schematically in Figure 2, panel C.

**TABLE II.**
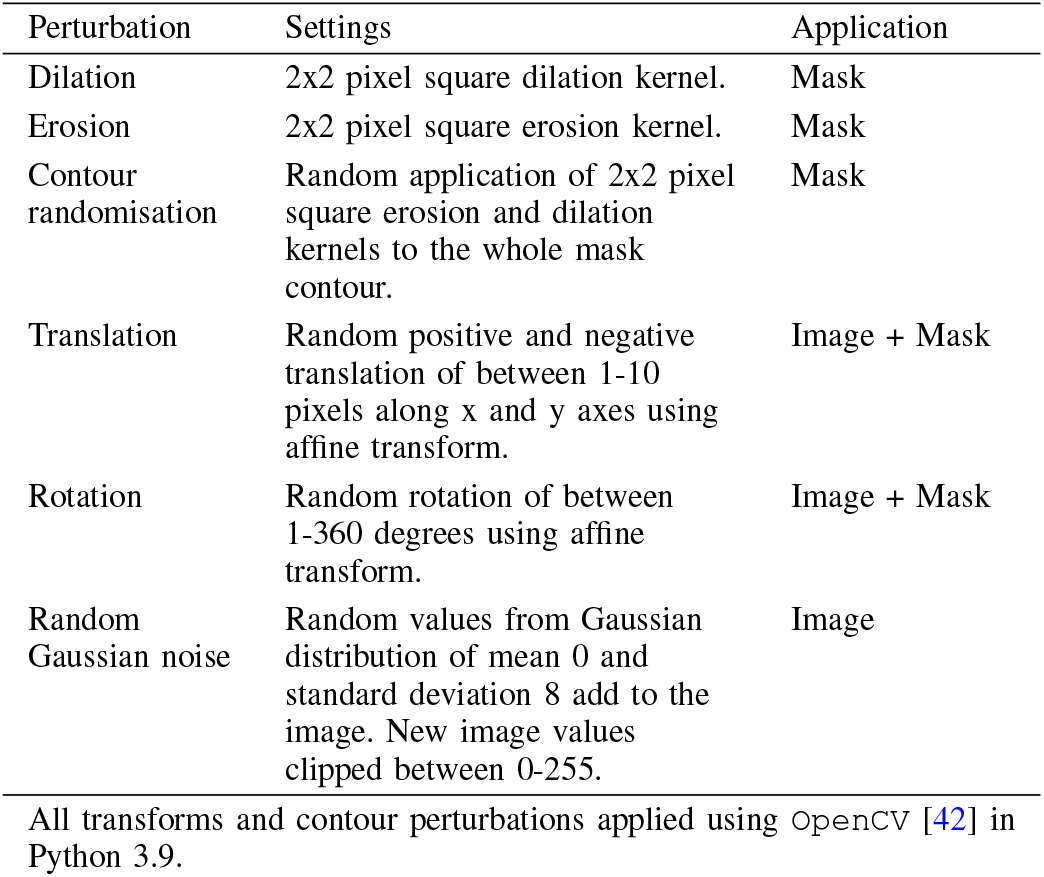
Image and mask perturbation settings.

| Perturbation | Settings | Application |
| --- | --- | --- |
| Dilation | 2x2 pixel square dilation kernel. | Mask |
| Erosion | 2x2 pixel square erosion kernel. | Mask |
| Contour randomisation | Random application of 2x2 pixel square erosion and dilation kernels to the whole mask contour. | Mask |
| Translation | Random positive and negative translation of between 1-10 pixels along x and y axes using affine transform. | Image + Mask |
| Rotation | Random rotation of between 1-360 degrees using affine transform. | Image + Mask |
| Random Gaussian noise | Random values from Gaussian distribution of mean 0 and standard deviation 8 add to the image. New image values clipped between 0-255. | Image |

All transforms and contour perturbations applied using OpenCV [42] in Python 3.9.

**Fig. 2.**
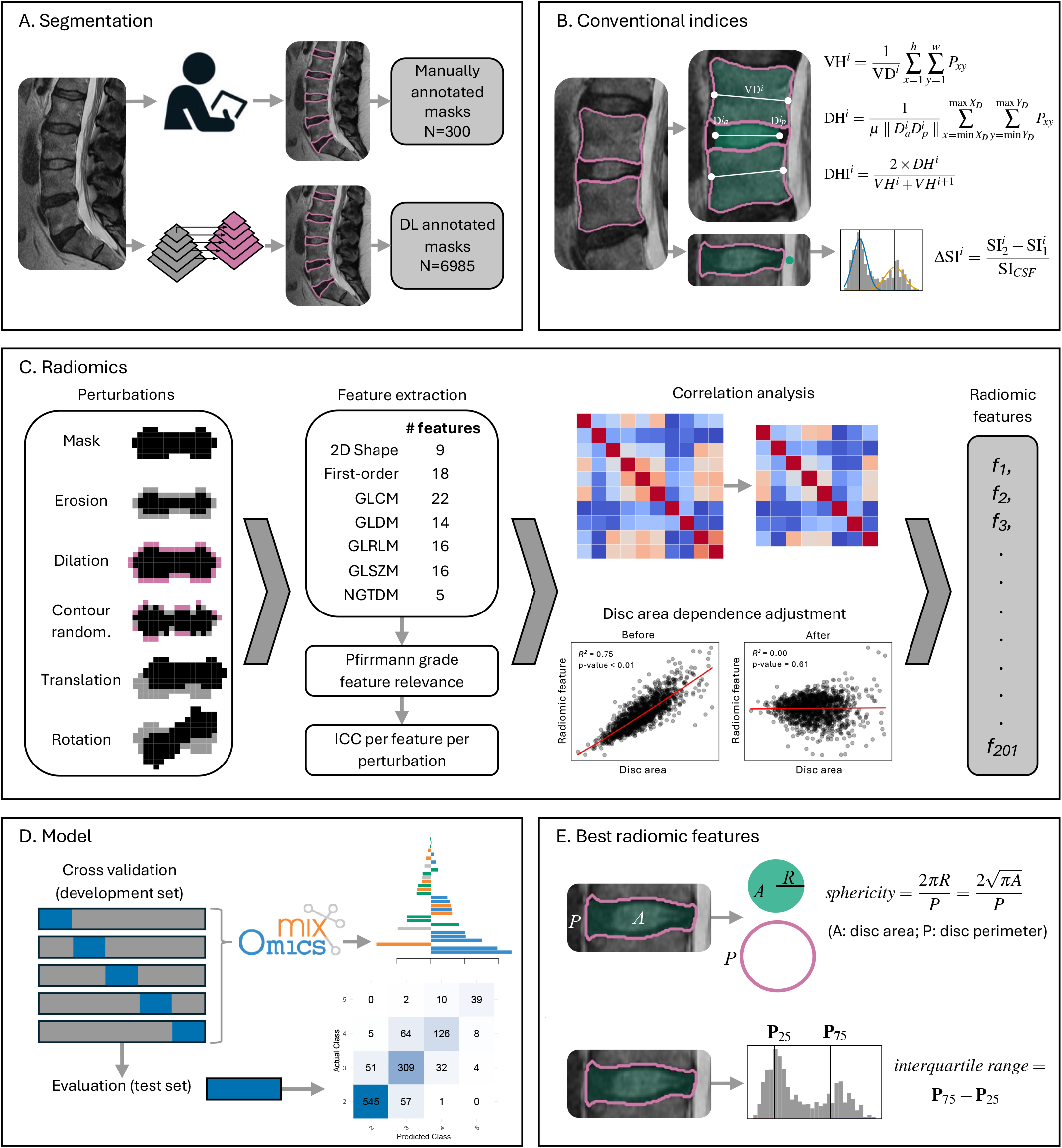
Overview of the methodological steps taken in the study: **A**. Deep learning and manual segmentation of the IVD and vertebral body; **B.** Calculation of the conventional indices per IVD; **C**. Radiomic feature calculation and robustness analysis; **D**. Classification model training and testing; **E**. Calculation of the two best performing radiomic features.

### B. Radiomics

After IVD segmentation and image-mask perturbations the workflow for the development of an IVD degeneration radiomic signature consisted of the following steps: 1) image preprocessing and extraction of radiomic features; 2) feature relevance, robustness, and correlation analysis; 3) IVD area-dependence adjustment; 4) model training and evaluation for Pfirrmann grade classification.

#### 1) Radiomic feature extraction

Features were calculated for all IVDs segmented by the DL model in the development and test datasets using the PyRadiomics package [43] and following the Imaging Biomarkers Initiative standards [44]. Due to anisotropy of T2-weighted lumbar spine sequences all features were calculated in 2D per slice and mean aggregated. The individual slices were normalized by centring gray values at the mean and scaling by the standard deviation after outlier removal [45]. Images were further pre-processed with discretisation of bin width 16 and two-fold resampling based on a preliminary analysis in the development set. The original (pre-processed) images were used, along with Laplacian of Gaussian filtered images with sigmas of 1, 3 and 5mm, and wavelet transformed images. For each set of images and matching IVD masks 18 first order intensity-based, 22 gray level co-occurrence (GLCM), 14 gray level dependence matrix (GLDM), 16 gray level run length matrix (GLRLM), 16 gray level size zone matrix (GLSZM), and 5 neighbouring gray tone difference matrix (NGTDM) features were calculated. From the masks 9 shape features were also calculated, resulting in a final set of 737 features per IVD. The same features were extracted from the 300 manually annotated IVDs and the 9 matched sets of image-mask perturbations.

#### 2) Feature relevance, robustness, and correlation analysis

In order to test the robustness of the the radiomic features, the intra-class correlation coefficient (ICC) for each feature for each perturbation was calculated against those calculated from the manual segmentations. The objective of selecting only the most robust features was balanced against the need to retain features of high relevance to Pfirrmann grade classification. To achieve this features with no statistically significant relationship with Pfirrmann grade were removed before carrying out a robustness analysis (Spearman’s correlation coefficient p-value *>* 0.05). Hierarchical clustering implemented in Scikit-learn [46] was then used to select a cluster of features that had high ICC values across perturbations. Details of this method are available in the supplementary material.

Correlation analysis was carried out to further reduce the number of features retained. Pairwise feature dependence was identified using Spearman’s correlation coefficient *>* 0.99. The feature with lower variance in each pair of highly correlated features was removed.

#### 3) IVD surface-area dependence adjustment

A number of radiomic features are volume or area dependent and correcting for this dependence has been shown to improve the performance of radiomics models [47], [48]. We tested the selected IVD features for their correlation to IVD 2D surface-area using linear, logarithmic, polynomial, power, and exponential models in the development set. The best fitting model for each feature was identified using information criterion and features with Spearman’s correlation coefficient of *>* 0.4 and p-value *<* 0.05 were then adjusted in the train and test sets using the models trained in the development set.

#### 4) Conventional indices

IVD height index was calculated using an area-based method [17], [18] as implemented by [20]. The IVD area was calculated from the middle 80% of the IVD mask and the height was obtained by dividing this area by its diameter. This was normalised to obtain the IVD height index by dividing by the sum of the adjacent vertebral body (VB) heights, which were also calculated using the area-based method (Figure 2, panel B.).

Peak signal intensity difference was calculated using the method of Waldenberg et al. [21] with the addition of a normalisation step. The peak signal intensity was calculated by fitting a two part Gaussian mixture model to the IVD signal intensity histogram and subtracting the respective peak values to obtain the absolute difference between the peaks (Figure 2, panel B). Higher values represent greater distinction between the darker annulus fibrosis voxels and lighter nucleus pulposis voxels. This was normalised for each IVD using the mean cerebrospinal fluid signal intensity from a single manually placed circular region of interest (ROI) of 3mm diameter placed as close as possible to the immediate posterior of the IVD in the slice with best visualisation of the spinal canal.

Further detail for the calculation of these features is available in the supplementary material. As with the radiomic features, 4 or 5 mid-sagittal slices were used and mean-aggregated. The ICCs for the conventional indices from manual vs DL segmentation were calculated in the same manner as the radiomic feature ICCs.

### C. Model training and evaluation

A sparse partial least squares discriminant analysis (sPLSDA) model was trained on the development set to classify the IVDs for Pfirrmann grade. sPLSDA was chosen due to the high dimensionality and multicollinearity of the radiomics data. The addition of sparsity simplified the model and facilitated some interpretation of the radiomic features. The model was implemented using the R package mixOmics [49]. Repeated 5-fold cross-validation was used to evaluate the model for the optimum number of components and feature sparsity per component based on balanced error rate. The final model trained in the development set was used to predict Pfirrmann grade in the test set. A second sPLSDA model was trained and tested using the same approach but excluding shape and first order features to examine and interpret the performance of texture features alone.

Feature importance was evaluated based on the sPLSDA loadings per component and via Spearman’s correlation coefficient for the correlation of individual features with Pfirrmann grade. Feature stability was evaluated as the proportion of cross validation folds (across repeats) where the feature was selected for a given component.

A support vector machine (SVM) was trained to classify Pfirrmann grade using IVD height index and peak signal intensity difference and evaluated on the test set. SVM hyper-parameters were tuned using repeated 5-fold cross validation in the development set.

## III. Results

### A. Data

#### 1) Segmentation reliability

Manual screening of the DL mask inferences resulted in the removal of lateral slices where the DL model failed. The most common reason for this was the presence of scoliosis in which the central region of the IVD or vertebral bodies that could be readily segmented was not captured by the numerical mid-sagittal slices. 5.6% of IVDs had 1-3 slices removed for this reason, but in all cases at least 1 accurately segmented mask per IVD could be retained for feature extraction.

Out of 300 manually annotated IVDs, the DL model misidentified the level in 17 cases. Excluding these, average agreement between the experienced musculoskeletal researcher (JM) and the DL segmentations was Dice coefficient 0.90 (0.87-0.94), Jaccard index 0.82 (0.77-0.88) and 95% Hausdorff of 2.12mm (0.60mm-3.65mm). The cases where the level was misidentified were also excluded for the radiomic feature robustness analysis.

#### 2) Feature relevance, reliability, and correlation analysis

307 features were removed because of low relevance to Pfirrmann grade (Spearman’s correlation coefficient p-value *>* 0.05. Clustering of the remaining features resulted in a set of 280 features with highest ICC values across perturbations. 79 features were highly correlated and removing those from each pair with less variance resulted in a final set of 201 features plus the two conventional indices that were retained for the Pfirrmann grade classification model.

ICC values and 95% confidence intervals for IVD height index and peak signal intensity difference were 0.83 (0.52-0.92) and 0.63 (0.17-0.81), respectively. Out of the 201 radiomic features retained, ICC ranged from 0.63-0.99, with 125 having an ICC greater than 0.9.

#### 3) IVD surface-area dependence adjustment

27 features were identified as highly correlated to IVD area and were adjusted in the development and test set using the coefficients of the model from the development set.

### B. Radiomic signature

#### 1) Classification model performance

The classification performance of the SVM model using conventional indices on the test set was balanced accuracy of 66.0% (62.0%-69.9% and Cohen’s Kappa of 0.55 (0.51-0.59). Repeated cross-validation resulted in a 24-component sPLSDA model using the texture features alone that achieved balanced accuracy of 72.5% (68.8%-76.1%) and Cohen’s Kappa of 0.66 (0.62-0.69) in the test set and a 23-component sPLSDA model using all radiomics features that achieved balanced accuracy of 76.8% (73.1%-80.3%) and Cohen’s Kappa of 0.71 (0.67-0.74) in the test set. The top features in both models were highly stable across cross-validation repeats. The full list of features used, their loadings, and stability is available in the supplementary material.

Given the high importance of two specific features in the full radiomics model, namely sphericity and interquartile range, a further SVM model was trained and tested using these two features with the same settings as the model for the conventional indices. This model classified the test set IVDs with BA of 76.7% (73.1%-80.1%) and Cohen’s Kappa of 0.67 (0.64-0.71). Evaluation metrics for all four models models are shown in Table III.

**TABLE III.**
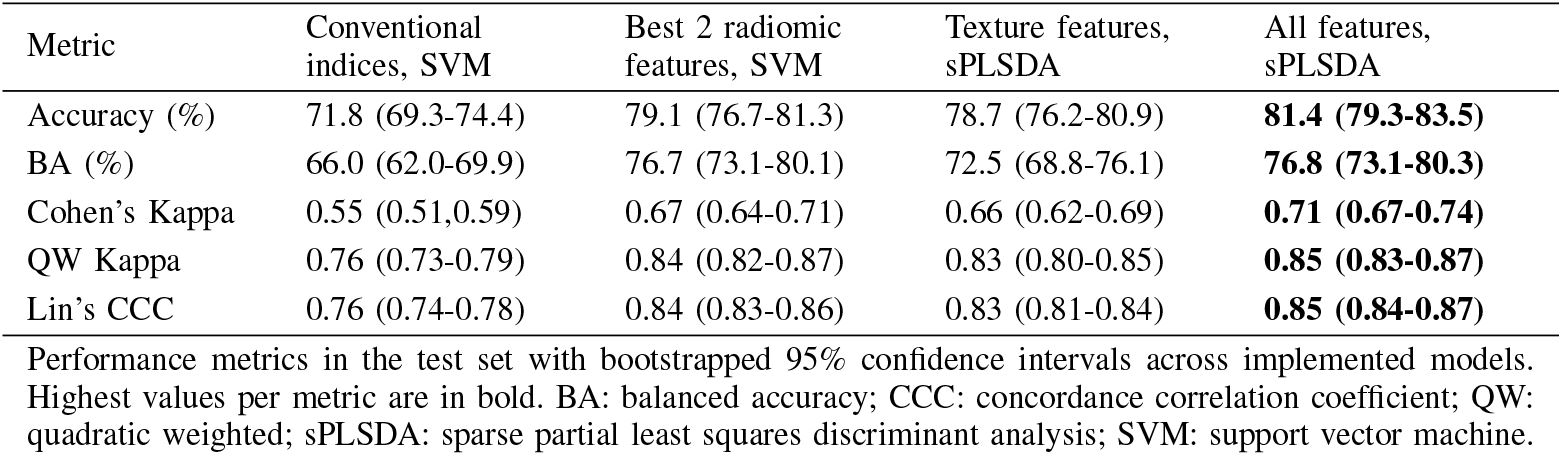
Pfirrmann grade classification model performance.

| Metric | Conventional indices, SVM | Best 2 radiomic features, SVM | Texture features, sPLSDA | All features, sPLSDA |
| --- | --- | --- | --- | --- |
| Accuracy (%) | 71.8 (69.3-74.4) | 79.1 (76.7-81.3) | 78.7 (76.2-80.9) | **81.4 (79.3-83.5)** |
| BA (%) | 66.0 (62.0-69.9) | 76.7 (73.1-80.1) | 72.5 (68.8-76.1) | **76.8 (73.1-80.3)** |
| Cohen's Kappa | 0.55 (0.51,0.59) | 0.67 (0.64-0.71) | 0.66 (0.62-0.69) | **0.71 (0.67-0.74)** |
| QW Kappa | 0.76 (0.73-0.79) | 0.84 (0.82-0.87) | 0.83 (0.80-0.85) | **0.85 (0.83-0.87)** |
| Lin's CCC | 0.76 (0.74-0.78) | 0.84 (0.83-0.86) | 0.83 (0.81-0.84) | **0.85 (0.84-0.87)** |

Performance metrics in the test set with bootstrapped 95% confidence intervals across implemented models. Highest values per metric are in bold. BA: balanced accuracy; CCC: concordance correlation coefficient; QW: quadratic weighted; sPLSDA: sparse partial least squares discriminant analysis; SVM: support vector machine.

Projections of the individual IVDs from the development set onto the first and second latent variables of the sPLSDA for the full radiomics model (panel A.) and the texture features model (panel B.) are shown in Figure 3. Scatter plots of the individual IVDs using the two conventional indices (panel A.) and the top-two radiomic features identified during the analysis (panel B.) are also shown in Figure 4 for comparison.

**Fig. 3.**
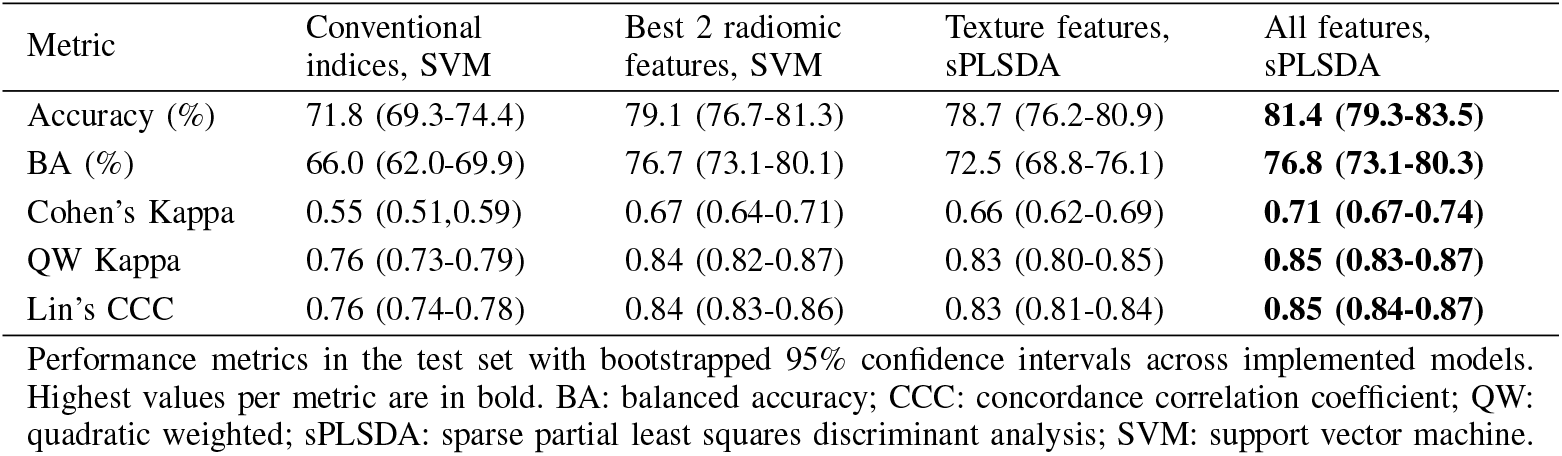
Scatter plots of the first two latent variables of the sPLSDA using all radiomic features (panel **A**.) and the sPLSDA using texture features only (panel **B**.). Ellipses represent 95% confidence for each Pfirrmann grade.

**Fig. 4.**
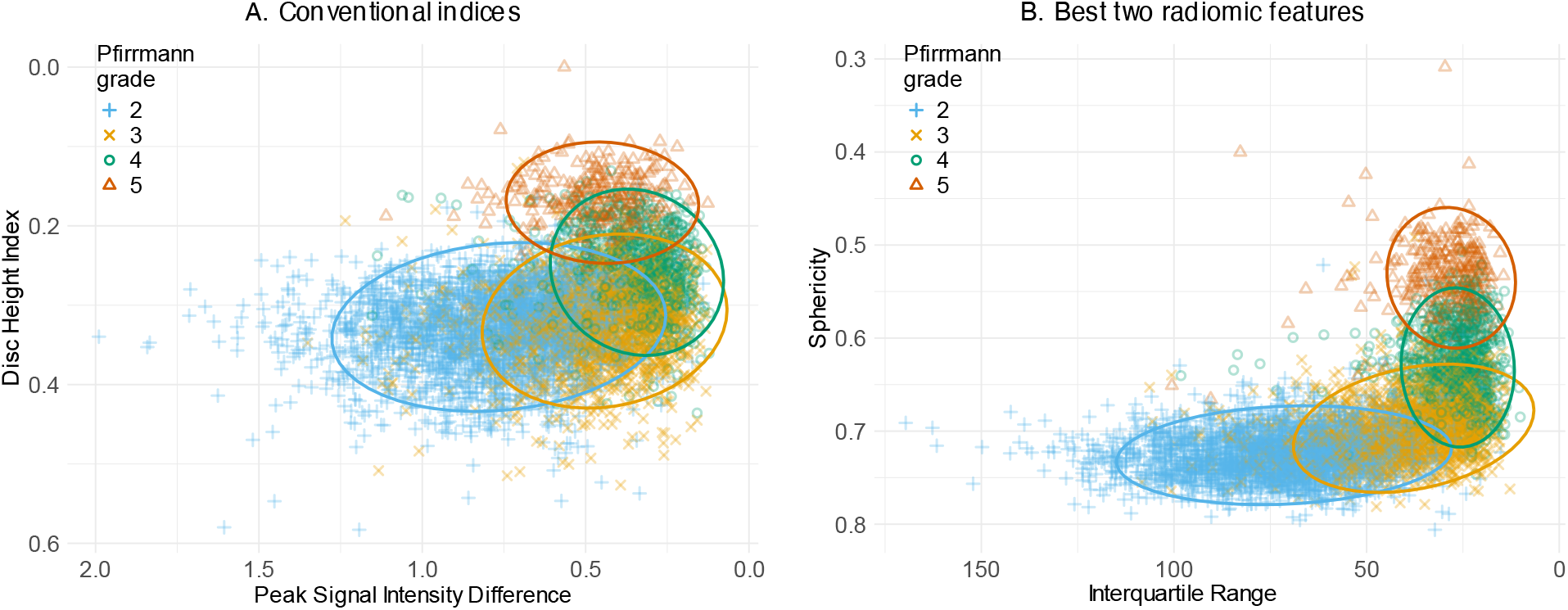
Scatter plots of the two conventional indices (panel **A**.) and the two radiomic features of highest importance from the sPLSDA model (panel **B**.). Ellipses represent 95% confidence for each Pfirrmann grade.

The 8 features contributing to the first two components of the full radiomics model are listed alongside the conventional indices in Table IV. Pfirrmann grade Spearman’s correlation coefficient was −0.67 (−0.65 − −0.68) for peak signal intensity difference and −0.45 (−0.43 − −0.47) for IVD height index, while ICC for these features from DL mask vs manual annotated mask was 0.60 and 0.81 respectively. Of the radiomic features with high importance, Spearman’s correlation coefficient was −0.77 (−0.76 − −0.78) for interquartile range, −0.72 (− 0.71 − − 0.73) for sphericity, and −0.69 (−0.68 − −0.71) for perimeter to surface ratio, with ICC values of 0.83, 0.96, and 0.96 respectively.

**TABLE IV.** Pfirrmann grade correlation and reliability of individual features.

| Feature | Correlation (95% CI) | ICC (95% CI) |
| --- | --- | --- |
| Conventional indices |  |  |
| Peak Signal Intensity Difference | -0.67 (-0.65 - -0.68) | 0.60 (0.22-0.78) |
| IVD height index | -0.45 (-0.43 - -0.47) | 0.81 (0.49-0.91) |
| All features sPLSDA component 1 |  |  |
| Interquartile Range | -0.77 (-0.76 - -0.78) | 0.83 (0.07-0.95) |
| All features sPLSDA component 2 |  |  |
| Sphericity | -0.72 (- 0.71 - -0.73) | 0.96 (0.870-0.98) |
| Perimeter to Surface Ratio | -0.69 (-0.68 - -0.71) | 0.96 (0.86-0.98) |
| LoG-3mm GLSZM Zone Percentage | 0.30 (0.28-0.32) | 0.97 (0.96-0.98) |
| LoG-3mm GLDM SDE | 0.17 (0.15-0.20) | 0.99 (0.98-0.99) |
| LoG-3mm GLCM Idn | -0.12 (-0.10 - -0.15) | 0.98 (0.97-0.99) |
| LoG-3mm GLRLM Run Percentage | -0.06 (-0.03 - -0.09) | 0.97 (0.13-0.99) |
| LoG-3mm GLRLM Run Length NUN | 0.05 (0.02-0.07) | 0.95 (0.07-0.99) |

Correlation column shows individual feature correlations with Pfirrmann grade using Spearman's correlation coefficient calculated on all deep learning IVD masks (development and test sets combined). ICC column shows ICC values for the features calculated from the deep learning IVD masks vs. the 283 manually annotated masks. All values have $p < 0.0001$ . CI: confidence interval; LoG: Laplacian of Gaussian; NUN: non-uniformity normalised; SDE: small dependence emphasis.

## IV. Discussion

In this study we identified a robust radiomic signature of IVD degeneration using DL segmentation that can effectively classify Pfirrmann grade and improves on the performance of conventional indices in the same data. The improvement is modest (65.99% vs 76.76% balanced accuracy, 0.55 vs 0.71 Cohen’s Kappa) but the radiomic features are more robust to IVD segmentation masks than the conventional indices. For example, the 8 features comprising the first two components of the sPLSDA latent space have ICC values ranging from 0.83-0.99, compared to IVD height index ICC of 0.81 and peak signal intensity difference ICC of 0.60. We additionally found that two individual radiomic features with the highest importance in the trained model could effectively classify Pfirrmann grade with comparable agreement and reliability to the full radiomics model (Table III). We propose these individual robust features, 2D sphericity and interquartile range (Figure 2, panel E.), as effective alternatives to the conventional indices of IVD height index and peak signal intensity difference for IVD degeneration classification.

Although the performance of our radiomics-based model did not match the performance state-of-the-art deep learning models trained and tested in the same dataset (e.g. [41] had balanced accuracy of 81.7% and Cohen’s Kappa of 0.74), the approach revealed new interpretable features of interest to IVD degeneration classification. For example, interquartile range may improve on peak signal intensity difference by reducing the influence of localized areas of high signal intensity associated with nitrogen gas or the gross morphological changes of severely degenerated IVDs. These can result in a U-shaped relationship between peak signal intensity difference and Pfirrmann grade where high values occur in both Pfirrmann grade 2 and grade 5 IVDs [15]. Sphericity may improve on IVD height index due to its ability to detect shape changes between grades 2 and 3, in contrast to IVD height index which remains constant between these grades. Sphericity may capture a combination of flattening and widening of the IVD and vertebral endplate disruption causing the perimeter to lengthen relative to the surface area (an aspect which is also captured by perimeter to surface ratio). Attempts have been made to incorporate this type of shape change into novel qualitative grading schemas [50] demonstrating the potential relevance of sphericity to degeneration classification.

Multiple studies have investigated other individual shape and textural descriptors for correlation with Pfirrmann grade. In one of the first studies of IVD degeneration texture features from T2-weighted MRI, Michopoulou et al. [31] found first order standard deviation had a Spearman’s correlation coefficient with Pfirrmann grade of −0.711, and GLCM sum of squares −0.812. Nagashima et al. [51] detailed an IVD signal intensity method involving manual placement of ROIs that had a correlation of −0.78 with Pfirrmann grade. Ruiz-Espana et al [30] derived intensity profiles from the IVD using a semi-automatic method and their features had Cohen’s K of 0.64-0.81 for Pfirrmann grade classification. Manual IVD height and signal intensity methods employed by Salamat et al [52] only found IVD height to effectively differentiate Pfirrmann grades 4 and 5, while their signal intensity method could differentiate Pfirrmann grades 1-4 but not 4 from 5. Yang et al. investigated the relative position of the nucleus pulposis along the longitudinal axis of the IVD for its relationship with Pfirrmann grades 1, 2, and 3 using manual annotation of the IVD and nucelus pulposis with statistically significant differences between the grades. Jarman et al. [25] found their IVD height index method to have statistically significant differences between the Pfirrmann grades, and Waldenberg et al.’s [21] peak signal intensity difference was statistically significant different across Pfirrmann grades.

While some of these features showed strong correlation with Pfirrmann grade, the studies are all characterised by small numbers of participants (*<* 100) and rely on manual or semi-automatic methods for IVD segmentation or ROI placement. The sensitivity of features to variations in the mask has not been explored despite the significant influence that segmentation variations can have on feature calculation and classification accuracy [54]. The only study we are aware of to examine the conventional indices from DL segmentations in T2-weighted MRI is Zheng et al. [20]. They used a DL IVD and VB segmentation model that had Dice scores ranging from 0.86-0.95 for the IVD masks and 0.92-0.97 for the VB masks compared to manual annotations, which represents similar performance to the DL segmentation model we used. They compared IVD height index derived automatically from the DL segmentations to a fully manual method carried out by orthopaedic residents, which resulted in an ICC of 0.90 for IVD height index. A similar comparison for peak signal intensity was not provided. Spearman’s correlation coefficients for IVD height index against their modified 8 point Pfirrmann grade ranged from −0.24 to −0.67 varying based on level and gender, while they report a single Spearman’s correlation coefficient of −0.96 for peak signal intensity difference across levels and both genders. While it is difficult to compare our study to these results directly, they indicate that even with accurate DL segmentations, IVD height index reliability and correlation with Pfirrmann grade is susceptible to significant variation when comparing manual and automated methods.

To summarise, few studies have examined radiomic features from T2-weighted MRI specifically in relation to Pfirrmann grade and we are not aware of other studies that have examined the impact of DL segmentation on these features. The studies that have been carried out lack detail on feature robustness and reliability, and pre-processing steps. The IVD surface area dependence of features that we identified has also not been considered by previous studies.

Although we corrected for IVD area dependence, visual inspection of the most important features (Figures 3 and 4) suggests that the information contained in the radiomic signature overlaps substantially with IVD height index and peak signal intensity difference. This overlap is further demonstrated by the observation that the two best radiomic features alone perform comparably to the more complex 23-component model using multiple features. This is unsurprising given the importance of IVD height and signal intensity in the Pfirrmann grading scale, compared to homogeneity or other qualitative imaging features of IVD degeneration. Future studies could explore texture features from standard-of-care MRI for clusters of IVDs that appear otherwise healthy (i.e., Pfirrmann grade 2) in an unsupervised approach, with the aim of identifying early sub-visual changes indicative of biochemical alterations known to precede visible degeneration [16].

There are also some limitations in this study. Firstly, we used numerical slice ordering to identify mid-sagittal slices, which may not align with the anatomical midline, especially in cases of scoliosis. Although manual screening helped mitigate the issue, future work should incorporate an optimization step to select slices closest to the anatomical midline. Secondly, additional aggregation methods (e.g., using the minimum, maximum, or range of feature values rather than the mean) warrant further exploration. Thirdly, accurate identification of IVD levels remains challenging for DL models, particularly in the presence of transitional vertebrae, and is likely to be a significant source of error in the model due to mismatching spinal levels. Transitional vertebrae have a prevalence of 16% in the NFBC1966, and in our validation of the DL segmentation model, over 5% of IVDs were labelled with the incorrect level. Identification of the anatomical midline per IVD, aggregation of data across slices, and transitional vertebrae all represent sources of error that likely impacted the performance of the model.

## V. Conclusion

Our results identify 2D sphericity and interquartile range as robust stand-alone texture features that are highly associated with Pfirrmann grade. These features, if validated, could serve as more reliable alternatives to the conventional indices of IVD height and peak signal intensity difference derived from DL segmentation. Although a more complex radiomic signature also accurately classifies Pfirrmann grade, it does not provide additional information beyond that captured by these key features. Overall, the robust radiomic features identified in this study represent a significant advance in the automated quantitative phenotyping of IVD degeneration from standard-of-care MRI. This approach has important implications for the measurement of IVD degeneration severity, monitoring of longitudinal change in the IVD, and prediction of IVD degeneration trajectories or treatment response in large clinical datasets and population cohorts.

## Data Availability

All data are available on request from the Northern Finland Birth Cohort 1966.

http://urn.fi/urn:nbn:fi:att:bc1e5408-980e-4a62-b899-43bec3755243

## VI. Acknowledgment

This research is part of a project that has received funding from the European Union’s Horizon 2020 research and innovation programme under the Marie Skłodowska-Curie grant agreement number 955735. This content reflects only the author’s view. It does not represent the view of the European Commission and the European Commission are not responsible for any use that may be made of the information it contains.

This research also received funding from the Finnish Cultural Foundation grant number 00240840.

We thank all NFBC1966 members and researchers who participated in the 46-year study. We also wish to acknowledge the work of the NFBC project centre.

