## Supplementary material for "Robust radiomic signatures of intervertebral disc degeneration from MRI"

### Calculation of conventional indices

Intervertebral disc (IVD) height index was calculated using an area-based method [2, 1] as implemented by [4]. The diameters of the two vertebral bodies (VBs) adjacent to each IVD were first determined (Equation 1). The four corners of the VB were identified using the Shi-Tomasi corner detection method. The vertebral body diameter ($\mathrm{VD}^i$) was calculated as the Euclidean distance between the midpoint of the line connecting the two anterior corners ($L^i_{\mathrm{amp},j}$) and the midpoint of the line connecting the two posterior corners ($L^i_{\mathrm{pmp},j}$):

$$\mathrm{VD}^i = \sqrt{\sum_{j=1}^{2}\left(L^i_{\mathrm{amp},j} - L^i_{\mathrm{pmp},j}\right)^2} \tag{1}$$

The vertebral height ($\mathrm{VH}^i$) was calculated as the surface area of the vertebral body divided by its diameter (Equation 2):

$$\mathrm{VH}^i = \frac{1}{\mathrm{VD}^i}\sum_{x=1}^{h}\sum_{y=1}^{w} P_{xy} \tag{2}$$

The IVD height was estimated using an area-based method, following Videman's approach. The IVD area was calculated from the central 80% of the IVD, and the height ($\mathrm{DH}^i$) was obtained by dividing the area by the IVD diameter (Equation 3):

$$\mathrm{DH}^i = \frac{1}{\mu \parallel D^i_a D^i_p \parallel}\sum_{x=\min X_D}^{\max X_D}\sum_{y=\min Y_D}^{\max Y_D} P_{xy} \tag{3}$$

The IVD height was normalized by dividing it by the sum of the heights of the two adjacent vertebrae, as shown in Equation 4:

$$\mathrm{DHI}^i = \frac{2 \times DH^i}{VH^i + VH^{i+1}} \tag{4}$$

The peak signal intensity (SI) difference was calculated using the method of Waldenberg [3], with an additional normalization step (Equation 5). The peak SI was determined by fitting a two-part Gaussian to the disc signal intensity histogram and subtracting the respective Gaussian peak values. Higher values indicated greater contrast between the darker annulus fibrosus (AF) and the lighter nucleus pulposus (NP). This value was normalized by dividing by the mean signal intensity of cerebrospinal fluid (CSF), measured from a circular region of interest (ROI) in the anterior dural sac (3mm diameter), placed as close as possible to the posterior aspect of the disc:

$$\Delta\mathrm{SI}^i = \frac{\mathrm{SI}^i_2 - \mathrm{SI}^i_1}{\mathrm{SI}_{CSF}} \tag{5}$$

### Calculation of best 2 radiomic features

Iterquartile range and 2D sphericity were identified as highly relevant radiomic features. These features were calculated using Pyradiomics. *sphericity* is given by:

$$sphericity = \frac{2\pi R}{P} = \frac{2\sqrt{\pi A}}{P} \tag{6}$$

Where $R$ is the radius of a circle with the same surface as the disc mask, and equal to $\sqrt{\frac{A}{\pi}}$, and $P$ is the length of the perimeter of the disc mask. Interquartile range is given by:

$$interquartile\ range = \mathbf{P}_{75} - \mathbf{P}_{25} \tag{7}$$

Where $\mathbf{P}_{25}$ and $\mathbf{P}_{75}$ are the 25$^{\text{th}}$ and 75$^{\text{th}}$ percentiles of the image array, respectively. All radiomic features other than 2D shape features were calculated on normalised images using the Pyradiomics package. Pyradiomics normalizes the image by centering it at the mean with standard deviation. Normalization uses all gray values in the image.

$$f(x) = \frac{x - \mu_x}{\sigma_x} \tag{8}$$

Where $x$ and $f(x)$ are the original and normalized intensity, respectively, and $\mu_x$ and $\sigma_x$ are the mean and standard deviation of the image intensity values. Using Pyradiomics, outliers in the image with values for which $x > \mu_x + 3\sigma_x$ or $x < \mu_x - 3\sigma_x$ were set to $\mu_x + 3\sigma_x$ and $\mu_x - 3\sigma_x$, respectively.

### Identification of and adjustment for disc area dependence

IVD area dependence was checked for all features by fitting linear, logarithmic, exponential, power, and polynomial models to each feature using IVD surface area as the independent variable for all IVDs in the development set. AIC and BIC were used to identify models with the best fit and features with R2 of > 0.4 and p-value of < 0.05 were considered highly area dependent and selected for adjustment. 27 features met these criteria and are listed in Table 1.

**Table 1.** IVD area dependent radiomic features and best fitting models used for feature adjustment.

| Feature | $R^2$ | Model |
|---|---|---|
| wavelet-LH_GrayLevelNonUniformity | 0.99 | power |
| wavelet-LH_GrayLevelNonUniformity | 0.89 | power |
| log-sigma-1-mm_GrayLevelNonUniformity | 0.87 | power |
| wavelet-LL_DependenceNonUniformity | 0.87 | power |
| wavelet-HL_RunLengthNonUniformity | 0.83 | power |
| wavelet-LH_DependenceNonUniformity | 0.82 | power |
| MinorAxisLength | 0.80 | power |
| log-sigma-5-mm_DependenceNonUniformity | 0.79 | power |
| TotalEnergy | 0.75 | power |
| log-sigma-1-mm_DependenceNonUniformity | 0.72 | power |
| log-sigma-1-mm_Coarseness | 0.70 | polynomial |
| log-sigma-3-mm_DependenceNonUniformity | 0.70 | power |
| PerimeterSurfaceRatio | 0.69 | polynomial |
| wavelet-HH_LargeAreaLowGrayLevelEmphasis | 0.69 | power |
| log-sigma-3-mm_LargeAreaHighGrayLevelEmphasis | 0.62 | power |
| log-sigma-5-mm_LargeAreaHighGrayLevelEmphasis | 0.55 | power |
| wavelet-LH_ZoneVariance | 0.53 | power |
| log-sigma-1-mm_GrayLevelNonUniformity | 0.51 | power |
| wavelet-HH_ZonePercentage | 0.49 | polynomial |
| log-sigma-5-mm_GrayLevelNonUniformity | 0.46 | power |
| log-sigma-5-mm_RunEntropy | 0.46 | power |
| log-sigma-3-mm_RunEntropy | 0.44 | power |
| log-sigma-1-mm_Median | 0.43 | power |
| log-sigma-1-mm_LargeAreaHighGrayLevelEmphasis | 0.43 | power |
| log-sigma-5-mm_LongRunHighGrayLevelEmphasis | 0.41 | power |
| log-sigma-5-mm_ZoneVariance | 0.40 | power |

*P-values of < 0.0001 for all listed features.*

Features were adjusted in the development set and the test set using the models trained on the development set alone to prevent data leakage. Examples of area dependent features plotted against IVD surface area are shown for the test set in Figure 1. The model predictions pre-adjustment are shown alongside the adjusted features with a new line of best fit.

### Feature robustness to image and mask perturbations

ICC2 values were calculated for each feature for each image and mask perturbation against the features calculated from the manually segmented discs. The relative impact on ICC values per perturbation is shown in 2. The feature ICC values across perturbations were clustered to identify a subset of features most robust to the image mask variations (3).

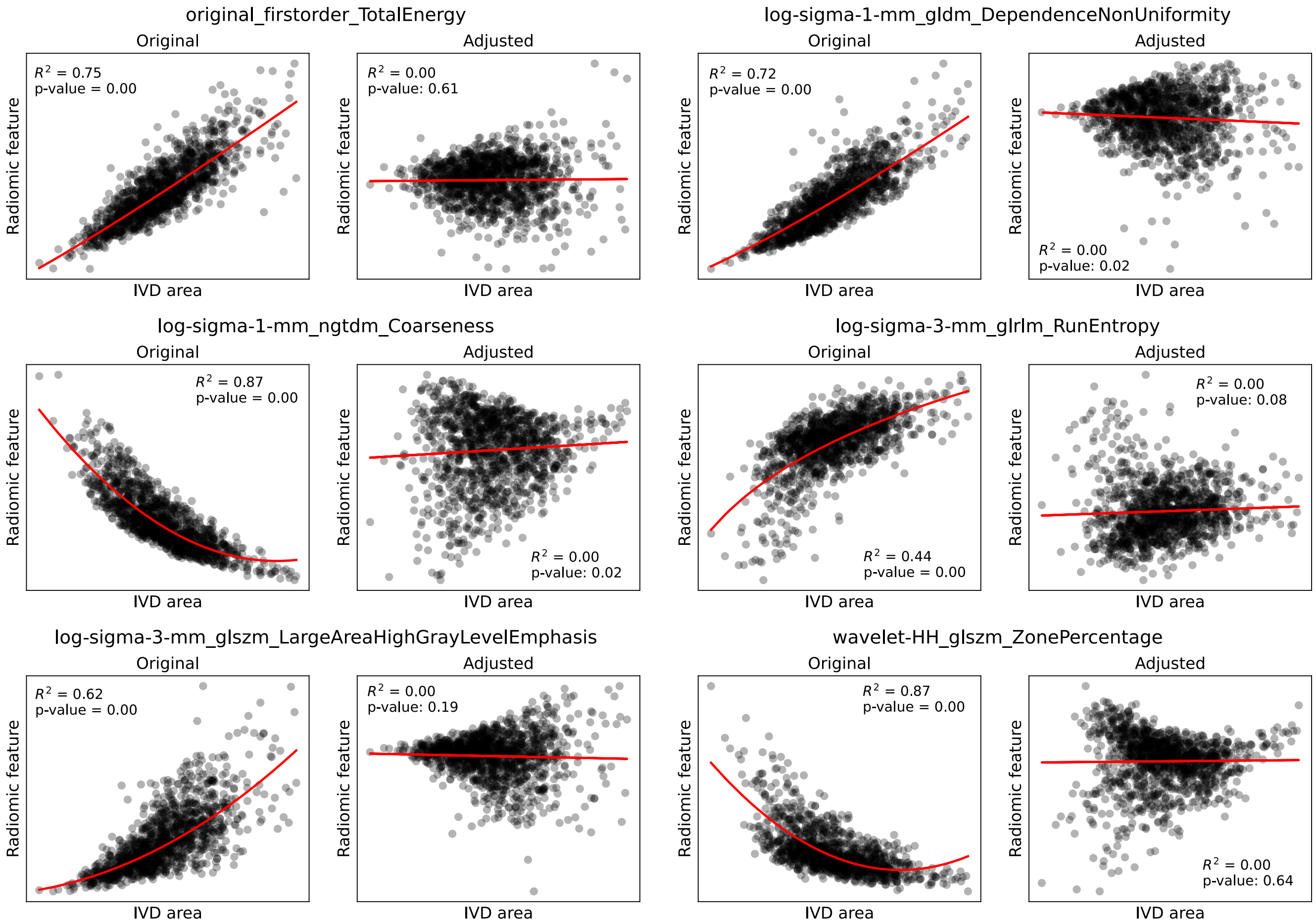


**Figure 1.** Examples of 6 IVD area dependent radiomic features from the test data plotted against IVD surface area. Panels labelled "original" show the features before area adjustment with a line of best fit in red, panels labelled "adjusted" show the features after adjustment for IVD area with a new line of best fit. The area dependence was adjusted independently in the development and test datasets.

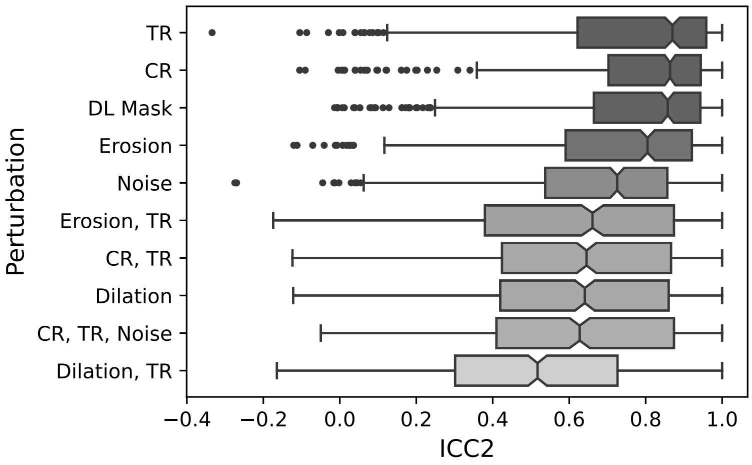


**Figure 2.** ICC2 values for all features for each image-mask perturbation, in descending order of median value per perturbation. TR: translation, rotation; CR: contour randomisation; DL: deep learning.

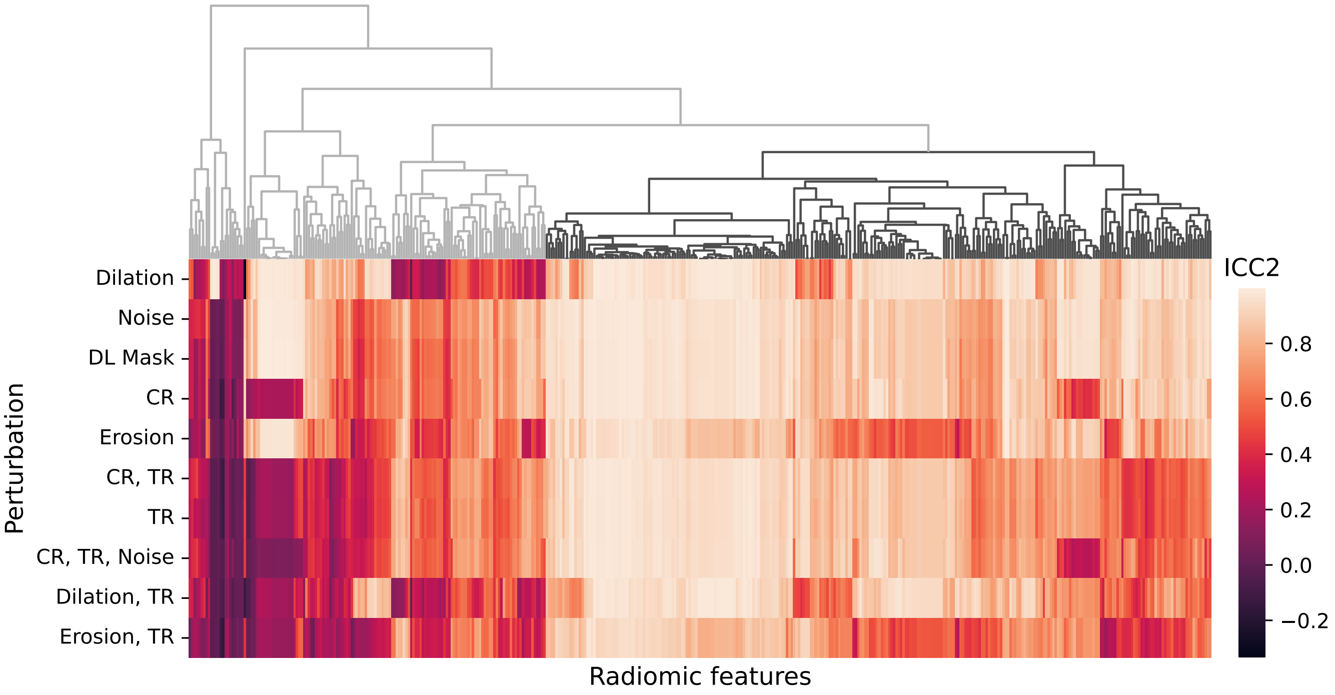


**Figure 3.** Hierarchically clustered heatmap of ICC2 values for 430 radiomic features (columns) for each image-mask perturbation (rows). The cluster dendrogram of 280 most robust features is shaded in dark gray, and the 4 remaining clusters of less robust features are shaded light gray. TR: translation, rotation; CR: contour randomisation; DL: deep learning.

**Table 2. List of final set of features used in alphabetical order:**

LoG-1mm_FirstOrder_90Percentile
LoG-1mm_FirstOrder_Mean
LoG-1mm_FirstOrder_Median
LoG-1mm_GLCM_DifferenceVariance
LoG-1mm_GLDM_DependenceNonUniformity
LoG-1mm_GLDM_DependenceNonUniformityNormalized
LoG-1mm_GLDM_DependenceVariance
LoG-1mm_GLDM_GrayLevelNonUniformity
LoG-1mm_GLDM_SmallDependenceEmphasis
LoG-1mm_GLRLM_GrayLevelNonUniformity
LoG-1mm_GLRLM_LongRunEmphasis
LoG-1mm_GLRLM_RunLengthNonUniformityNormalized
LoG-1mm_GLRLM_RunPercentage
LoG-1mm_GLRLM_RunVariance
LoG-1mm_GLRLM_ShortRunEmphasis
LoG-1mm_GLSZM_LargeAreaEmphasis
LoG-1mm_GLSZM_LargeAreaHighGrayLevelEmphasis
LoG-1mm_GLSZM_LargeAreaLowGrayLevelEmphasis
LoG-1mm_GLSZM_ZonePercentage
LoG-1mm_NGTDM_Coarseness
LoG-1mm_NGTDM_Strength
LoG-3mm_FirstOrder_10Percentile
LoG-3mm_FirstOrder_90Percentile
LoG-3mm_FirstOrder_Entropy
LoG-3mm_FirstOrder_InterquartileRange
LoG-3mm_FirstOrder_Kurtosis
LoG-3mm_FirstOrder_Maximum
LoG-3mm_FirstOrder_MeanAbsoluteDeviation
LoG-3mm_FirstOrder_Median
LoG-3mm_FirstOrder_RootMeanSquared
LoG-3mm_FirstOrder_Skewness
LoG-3mm_FirstOrder_Uniformity
LoG-3mm_GLCM_ClusterShade
LoG-3mm_GLCM_Correlation
LoG-3mm_GLCM_DifferenceEntropy
LoG-3mm_GLCM_Idmn
LoG-3mm_GLCM_Idn
LoG-3mm_GLCM_Imc1
LoG-3mm_GLCM_JointEntropy
LoG-3mm_GLCM_MaximumProbability
LoG-3mm_GLDM_DependenceEntropy
LoG-3mm_GLDM_DependenceNonUniformity

LoG-3mm_GLDM_DependenceNonUniformityNormalized
LoG-3mm_GLDM_DependenceVariance
LoG-3mm_GLDM_GrayLevelNonUniformity
LoG-3mm_GLDM_SmallDependenceEmphasis
LoG-3mm_GLDM_SmallDependenceHighGrayLevelEmphasis
LoG-3mm_GLRLM_GrayLevelNonUniformity
LoG-3mm_GLRLM_LongRunEmphasis
LoG-3mm_GLRLM_LongRunLowGrayLevelEmphasis
LoG-3mm_GLRLM_RunEntropy
LoG-3mm_GLRLM_RunLengthNonUniformity
LoG-3mm_GLRLM_RunLengthNonUniformityNormalized
LoG-3mm_GLRLM_RunPercentage
LoG-3mm_GLRLM_RunVariance
LoG-3mm_GLRLM_ShortRunEmphasis
LoG-3mm_GLRLM_ShortRunHighGrayLevelEmphasis
LoG-3mm_GLSZM_LargeAreaHighGrayLevelEmphasis
LoG-3mm_GLSZM_LargeAreaLowGrayLevelEmphasis
LoG-3mm_GLSZM_SizeZoneNonUniformity
LoG-3mm_GLSZM_ZonePercentage
LoG-3mm_GLSZM_ZoneVariance
LoG-3mm_NGTDM_Complexity
LoG-3mm_NGTDM_Contrast
LoG-5mm_FirstOrder_10Percentile
LoG-5mm_FirstOrder_90Percentile
LoG-5mm_FirstOrder_Kurtosis
LoG-5mm_FirstOrder_Median
LoG-5mm_FirstOrder_RootMeanSquared
LoG-5mm_FirstOrder_Skewness
LoG-5mm_FirstOrder_TotalEnergy
LoG-5mm_GLCM_ClusterShade
LoG-5mm_GLCM_Correlation
LoG-5mm_GLCM_DifferenceAverage
LoG-5mm_GLCM_Idmn
LoG-5mm_GLCM_Idn
LoG-5mm_GLCM_Imc1
LoG-5mm_GLCM_JointAverage
LoG-5mm_GLDM_DependenceEntropy
LoG-5mm_GLDM_DependenceNonUniformity
LoG-5mm_GLDM_DependenceVariance
LoG-5mm_GLDM_GrayLevelNonUniformity
LoG-5mm_GLDM_LargeDependenceLowGrayLevelEmphasis
LoG-5mm_GLDM_SmallDependenceEmphasis
LoG-5mm_GLDM_SmallDependenceHighGrayLevelEmphasis

LoG-5mm_GLRLM_LongRunEmphasis
LoG-5mm_GLRLM_LongRunHighGrayLevelEmphasis
LoG-5mm_GLRLM_LongRunLowGrayLevelEmphasis
LoG-5mm_GLRLM_RunEntropy
LoG-5mm_GLRLM_RunLengthNonUniformity
LoG-5mm_GLRLM_RunLengthNonUniformityNormalized
LoG-5mm_GLRLM_RunPercentage
LoG-5mm_GLRLM_RunVariance
LoG-5mm_GLRLM_ShortRunEmphasis
LoG-5mm_GLRLM_ShortRunHighGrayLevelEmphasis
LoG-5mm_GLSZM_GrayLevelNonUniformity
LoG-5mm_GLSZM_LargeAreaEmphasis
LoG-5mm_GLSZM_LargeAreaHighGrayLevelEmphasis
LoG-5mm_GLSZM_LargeAreaLowGrayLevelEmphasis
LoG-5mm_GLSZM_SizeZoneNonUniformity
LoG-5mm_GLSZM_ZoneEntropy
LoG-5mm_GLSZM_ZonePercentage
LoG-5mm_GLSZM_ZoneVariance
LoG-5mm_NGTDM_Contrast
Original_Elongation
Original_FirstOrder_10Percentile
Original_FirstOrder_InterquartileRange
Original_FirstOrder_TotalEnergy
Original_FirstOrder_Uniformity
Original_GLCM_JointAverage
Original_GLCM_JointEnergy
Original_GLCM_MaximumProbability
Original_GLDM_DependenceNonUniformity
Original_GLDM_DependenceVariance
Original_GLDM_GrayLevelNonUniformity
Original_GLDM_LargeDependenceHighGrayLevelEmphasis
Original_GLDM_LargeDependenceLowGrayLevelEmphasis
Original_GLDM_LowGrayLevelEmphasis
Original_GLDM_SmallDependenceLowGrayLevelEmphasis
Original_GLRLM_GrayLevelNonUniformity
Original_GLRLM_LongRunHighGrayLevelEmphasis
Original_GLRLM_LongRunLowGrayLevelEmphasis
Original_GLSZM_LargeAreaEmphasis
Original_GLSZM_LargeAreaHighGrayLevelEmphasis
Original_GLSZM_SizeZoneNonUniformityNormalized
Original_MajorAxisLength
Original_MaximumDiameter
Original_MinorAxisLength

Original_PerimeterSurfaceRatio
Original_Sphericity
Wavelet_HH_FirstOrder_Variance
Wavelet_HH_GLDM_SmallDependenceEmphasis
Wavelet_HH_GLRLM_GrayLevelNonUniformity
Wavelet_HH_GLRLM_RunLengthNonUniformity
Wavelet_HH_GLSZM_LargeAreaHighGrayLevelEmphasis
Wavelet_HH_GLSZM_LargeAreaLowGrayLevelEmphasis
Wavelet_HH_GLSZM_ZonePercentage
Wavelet_HH_GLSZM_ZoneVariance
Wavelet_HL_FirstOrder_Kurtosis
Wavelet_HL_FirstOrder_Maximum
Wavelet_HL_FirstOrder_Range
Wavelet_HL_GLCM_DifferenceAverage
Wavelet_HL_GLCM_Idm
Wavelet_HL_GLCM_InverseVariance
Wavelet_HL_GLCM_MaximumProbability
Wavelet_HL_GLDM_DependenceEntropy
Wavelet_HL_GLDM_DependenceNonUniformity
Wavelet_HL_GLDM_DependenceNonUniformityNormalized
Wavelet_HL_GLDM_DependenceVariance
Wavelet_HL_GLDM_SmallDependenceEmphasis
Wavelet_HL_GLRLM_GrayLevelNonUniformity
Wavelet_HL_GLRLM_LongRunEmphasis
Wavelet_HL_GLRLM_RunLengthNonUniformity
Wavelet_HL_GLRLM_RunLengthNonUniformityNormalized
Wavelet_HL_GLRLM_RunVariance
Wavelet_HL_GLRLM_ShortRunEmphasis
Wavelet_HL_GLSZM_LargeAreaHighGrayLevelEmphasis
Wavelet_HL_GLSZM_ZonePercentage
Wavelet_HL_NGTDM_Complexity
Wavelet_LH_FirstOrder_10Percentile
Wavelet_LH_FirstOrder_Maximum
Wavelet_LH_FirstOrder_Range
Wavelet_LH_GLCM_DifferenceAverage
Wavelet_LH_GLCM_Imc1
Wavelet_LH_GLDM_DependenceEntropy
Wavelet_LH_GLDM_DependenceNonUniformity
Wavelet_LH_GLDM_DependenceNonUniformityNormalized
Wavelet_LH_GLDM_DependenceVariance
Wavelet_LH_GLDM_GrayLevelNonUniformity
Wavelet_LH_GLDM_SmallDependenceEmphasis
Wavelet_LH_GLRLM_GrayLevelNonUniformity

| |
|---|
| Wavelet_LH_GLRLM_GrayLevelNonUniformityNormalized |
| Wavelet_LH_GLRLM_LongRunLowGrayLevelEmphasis |
| Wavelet_LH_GLRLM_RunEntropy |
| Wavelet_LH_GLRLM_RunLengthNonUniformityNormalized |
| Wavelet_LH_GLRLM_RunPercentage |
| Wavelet_LH_GLRLM_RunVariance |
| Wavelet_LH_GLRLM_ShortRunEmphasis |
| Wavelet_LH_GLSZM_LargeAreaHighGrayLevelEmphasis |
| Wavelet_LH_GLSZM_LargeAreaLowGrayLevelEmphasis |
| Wavelet_LH_GLSZM_ZonePercentage |
| Wavelet_LH_GLSZM_ZoneVariance |
| Wavelet_LH_NGTDM_Busyness |
| Wavelet_LH_NGTDM_Strength |
| Wavelet_LL_FirstOrder_Median |
| Wavelet_LL_FirstOrder_RootMeanSquared |
| Wavelet_LL_FirstOrder_TotalEnergy |
| Wavelet_LL_GLCM_InverseVariance |
| Wavelet_LL_GLCM_JointAverage |
| Wavelet_LL_GLCM_MaximumProbability |
| Wavelet_LL_GLDM_DependenceNonUniformity |
| Wavelet_LL_GLDM_DependenceNonUniformityNormalized |
| Wavelet_LL_GLDM_DependenceVariance |
| Wavelet_LL_GLDM_SmallDependenceLowGrayLevelEmphasis |
| Wavelet_LL_GLRLM_GrayLevelNonUniformity |
| Wavelet_LL_GLRLM_LongRunHighGrayLevelEmphasis |
| Wavelet_LL_GLRLM_LongRunLowGrayLevelEmphasis |
| Wavelet_LL_GLRLM_LowGrayLevelRunEmphasis |
| Wavelet_LL_GLSZM_GrayLevelNonUniformity |
| Wavelet_LL_GLSZM_LargeAreaHighGrayLevelEmphasis |
| Wavelet_LL_GLSZM_SmallAreaEmphasis |

**Table 3. Features per component, loadings, and stability (top 10 features shown per component where more than 10 were included)**

| Feature | Component | Class | Importance | Stability |
|---|---|---|---|---|
| Original_FirstOrder_InterquartileRange | 1 | 2 | -1.000 | 1 |
| LoG-3mm_GLCM_Idn | 2 | 3 | -0.236 | 1 |
| LoG-3mm_GLDM_SmallDependenceEmphasis | 2 | 5 | 0.632 | 1 |
| LoG-3mm_GLRLM_RunLengthNonUniformityNormalized | 2 | 5 | 0.201 | 1 |
| LoG-3mm_GLRLM_RunPercentage | 2 | 5 | 0.279 | 1 |
| LoG-3mm_GLSZM_ZonePercentage | 2 | 5 | 0.312 | 1 |
| Original_PerimeterSurfaceRatio | 2 | 2 | -0.245 | 1 |
| Original_Sphericity | 2 | 2 | -0.519 | 1 |

| | | | | |
|---|---|---|---|---|
| LoG-5mm_FirstOrder_TotalEnergy | 3 | 2 | -0.210 | 1 |
| LoG-5mm_GLCM_DifferenceAverage | 3 | 5 | 0.172 | 1 |
| LoG-5mm_GLCM_JointAverage | 3 | 5 | 0.189 | 1 |
| LoG-5mm_GLDM_DependenceNonUniformity | 3 | 5 | 0.208 | 1 |
| LoG-5mm_GLDM_LargeDependenceLowGrayLevelEmphasis | 3 | 2 | -0.159 | 1 |
| LoG-5mm_GLDM_SmallDependenceHighGrayLevelEmphasis | 3 | 5 | 0.162 | 1 |
| LoG-5mm_GLRLM_RunLengthNonUniformity | 3 | 5 | 0.210 | 1 |
| LoG-5mm_GLSZM_ZoneEntropy | 3 | 5 | 0.187 | 1 |
| Original_GLSZM_SizeZoneNonUniformityNormalized | 3 | 5 | 0.171 | 1 |
| Wavelet_HH_FirstOrder_Variance | 3 | 5 | 0.155 | 1 |
| Original_MajorAxisLength | 4 | 5 | 0.628 | 1 |
| Original_MaximumDiameter | 4 | 5 | 0.758 | 1 |
| Original_MinorAxisLength | 4 | 4 | 0.112 | 0.952 |
| Original_PerimeterSurfaceRatio | 4 | 2 | -0.117 | 0.88 |
| Wavelet_LL_GLDM_DependenceVariance | 4 | 5 | -0.066 | 0.96 |
| IVD_height_index | 5 | 2 | -0.205 | 1 |
| LoG-5mm_FirstOrder_Skewness | 5 | 3 | -0.235 | 1 |
| LoG-5mm_GLCM_Correlation | 5 | 2 | -0.221 | 1 |
| LoG-5mm_GLCM_Idmn | 5 | 2 | -0.220 | 1 |
| LoG-5mm_GLRLM_RunEntropy | 5 | 4 | 0.191 | 1 |
| Original_Elongation | 5 | 2 | -0.190 | 1 |
| Original_FirstOrder_Uniformity | 5 | 4 | 0.192 | 1 |
| Original_GLCM_JointEnergy | 5 | 4 | 0.196 | 1 |
| Original_GLCM_MaximumProbability | 5 | 4 | 0.197 | 1 |
| Wavelet_LL_GLCM_MaximumProbability | 5 | 5 | 0.167 | 1 |
| LoG-1mm_GLSZM_LargeAreaLowGrayLevelEmphasis | 6 | 3 | -0.323 | 1 |
| LoG-3mm_FirstOrder_Maximum | 6 | 5 | 0.168 | 1 |
| LoG-3mm_GLCM_ClusterShade | 6 | 2 | 0.598 | 1 |
| LoG-3mm_GLRLM_GrayLevelNonUniformity | 6 | 2 | 0.395 | 1 |
| Original_FirstOrder_10Percentile | 6 | 2 | 0.208 | 1 |
| Original_GLDM_DependenceNonUniformity | 6 | 2 | 0.205 | 0.984 |
| Original_GLDM_SmallDependenceLowGrayLevelEmphasis | 6 | 5 | -0.140 | 1 |
| Original_GLSZM_LargeAreaEmphasis | 6 | 4 | -0.195 | 1 |
| Wavelet_HH_FirstOrder_Variance | 6 | 5 | -0.140 | 0.768 |
| Wavelet_HH_GLDM_SmallDependenceEmphasis | 6 | 2 | 0.303 | 1 |
| LoG-1mm_GLRLM_GrayLevelNonUniformity | 7 | 2 | -0.096 | 0.28 |
| Original_MinorAxisLength | 7 | 4 | 0.995 | 0.32 |
| LoG-1mm_NGTDM_Strength | 8 | 5 | -0.207 | 0.256 |
| LoG-5mm_GLSZM_GrayLevelNonUniformity | 8 | 5 | 0.143 | 0.456 |
| Wavelet_LH_GLDM_DependenceVariance | 8 | 5 | -0.524 | 0.4 |
| Wavelet_LH_GLDM_SmallDependenceEmphasis | 8 | 5 | -0.246 | 0.416 |
| Wavelet_LH_GLRLM_GrayLevelNonUniformityNormalized | 8 | 3 | 0.243 | 0.384 |
| Wavelet_LH_GLRLM_LongRunLowGrayLevelEmphasis | 8 | 2 | 0.116 | 0.296 |

| | | | | |
|---|---|---|---|---|
| Wavelet_LH_GLRLM_ShortRunEmphasis | 8 | 5 | -0.182 | 0.4 |
| Wavelet_LH_GLSZM_LargeAreaLowGrayLevelEmphasis | 8 | 3 | 0.220 | 0.376 |
| Wavelet_LH_GLSZM_ZonePercentage | 8 | 5 | -0.654 | 0.416 |
| Wavelet_LH_NGTDM_Strength | 8 | 5 | -0.142 | 0.288 |
| Original_GLRLM_GrayLevelNonUniformity | 9 | 2 | -0.160 | 0.984 |
| Wavelet_HH_FirstOrder_Variance | 9 | 5 | 0.170 | 0.904 |
| Wavelet_HL_FirstOrder_Range | 9 | 3 | 0.218 | 0.824 |
| Wavelet_HL_GLCM_DifferenceAverage | 9 | 5 | 0.157 | 0.896 |
| Wavelet_HL_GLDM_SmallDependenceEmphasis | 9 | 5 | 0.168 | 0.928 |
| Wavelet_HL_NGTDM_Complexity | 9 | 3 | 0.190 | 0.824 |
| Wavelet_LH_FirstOrder_10Percentile | 9 | 4 | 0.162 | 1 |
| Wavelet_LH_GLDM_DependenceEntropy | 9 | 5 | 0.150 | 0.992 |
| Wavelet_LH_GLRLM_RunVariance | 9 | 2 | -0.191 | 0.968 |
| Wavelet_LL_GLRLM_GrayLevelNonUniformity | 9 | 4 | 0.163 | 0.952 |
| LoG-3mm_GLSZM_LargeAreaLowGrayLevelEmphasis | 10 | 3 | -0.153 | 0.992 |
| Wavelet_HH_GLDM_SmallDependenceEmphasis | 10 | 2 | -0.172 | 1 |
| Wavelet_HL_GLCM_DifferenceAverage | 10 | 5 | -0.174 | 1 |
| Wavelet_HL_GLCM_Idm | 10 | 2 | 0.184 | 1 |
| Wavelet_HL_GLCM_InverseVariance | 10 | 3 | -0.185 | 0.992 |
| Wavelet_HL_GLDM_DependenceEntropy | 10 | 5 | -0.164 | 1 |
| Wavelet_HL_GLRLM_LongRunEmphasis | 10 | 2 | 0.203 | 0.984 |
| Wavelet_HL_GLRLM_RunVariance | 10 | 2 | 0.165 | 1 |
| Wavelet_HL_GLRLM_ShortRunEmphasis | 10 | 5 | -0.167 | 1 |
| Wavelet_LH_GLRLM_GrayLevelNonUniformity | 10 | 2 | 0.151 | 0.984 |
| LoG-3mm_FirstOrder_Kurtosis | 11 | 3 | 0.188 | 0.92 |
| LoG-3mm_FirstOrder_Uniformity | 11 | 4 | 0.206 | 0.888 |
| LoG-3mm_GLCM_MaximumProbability | 11 | 4 | 0.202 | 0.88 |
| LoG-5mm_GLRLM_LongRunLowGrayLevelEmphasis | 11 | 2 | -0.175 | 0.88 |
| LoG-5mm_GLSZM_GrayLevelNonUniformity | 11 | 5 | -0.174 | 0.792 |
| LoG-5mm_GLSZM_SizeZoneNonUniformity | 11 | 5 | -0.186 | 0.904 |
| Original_FirstOrder_10Percentile | 11 | 2 | 0.192 | 0.856 |
| Wavelet_HH_GLRLM_GrayLevelNonUniformity | 11 | 5 | 0.169 | 0.856 |
| Wavelet_LL_GLDM_DependenceNonUniformity | 11 | 4 | -0.169 | 0.928 |
| Wavelet_LL_GLDM_DependenceNonUniformityNormalized | 11 | 2 | 0.166 | 0.856 |
| LoG-3mm_GLSZM_SizeZoneNonUniformity | 12 | 5 | -0.217 | 0.584 |
| LoG-5mm_GLSZM_SizeZoneNonUniformity | 12 | 5 | -0.272 | 0.8 |
| Wavelet_HH_GLRLM_GrayLevelNonUniformity | 12 | 5 | -0.273 | 0.696 |
| Wavelet_HH_GLRLM_RunLengthNonUniformity | 12 | 5 | -0.269 | 0.68 |
| Wavelet_HL_GLCM_Idm | 12 | 2 | -0.214 | 0.32 |
| Wavelet_HL_GLCM_InverseVariance | 12 | 3 | 0.254 | 0.336 |
| Wavelet_HL_GLDM_DependenceNonUniformity | 12 | 5 | -0.227 | 0.4 |
| Wavelet_HL_GLRLM_GrayLevelNonUniformity | 12 | 2 | -0.213 | 0.272 |
| Wavelet_HL_GLRLM_RunLengthNonUniformity | 12 | 2 | -0.285 | 0.344 |

| | | | | |
|---|---|---|---|---|
| Wavelet_HL_GLRLM_RunLengthNonUniformityNormalized | 12 | 5 | 0.356 | 0.272 |
| LoG-1mm_GLRLM_RunVariance | 13 | 3 | 0.202 | 0.752 |
| LoG-3mm_FirstOrder_InterquartileRange | 13 | 2 | 0.195 | 0.496 |
| LoG-5mm_GLRLM_LongRunHighGrayLevelEmphasis | 13 | 2 | 0.274 | 0.672 |
| LoG-5mm_GLSZM_LargeAreaHighGrayLevelEmphasis | 13 | 2 | 0.229 | 0.496 |
| LoG-5mm_GLSZM_SizeZoneNonUniformity | 13 | 5 | -0.305 | 0.712 |
| LoG-5mm_NGTDM_Contrast | 13 | 2 | 0.378 | 0.848 |
| Wavelet_HH_GLDM_SmallDependenceEmphasis | 13 | 2 | 0.213 | 0.672 |
| Wavelet_HH_GLRLM_GrayLevelNonUniformity | 13 | 5 | 0.222 | 0.608 |
| Wavelet_LH_GLDM_GrayLevelNonUniformity | 13 | 2 | 0.256 | 0.6 |
| Wavelet_LL_GLSZM_GrayLevelNonUniformity | 13 | 2 | 0.215 | 0.76 |
| LoG-1mm_GLDM_DependenceNonUniformityNormalized | 14 | 3 | -0.141 | 1 |
| LoG-1mm_GLRLM_GrayLevelNonUniformity | 14 | 2 | -0.282 | 0.984 |
| LoG-1mm_GLRLM_RunPercentage | 14 | 2 | 0.137 | 0.968 |
| Wavelet_HH_FirstOrder_Variance | 14 | 5 | 0.170 | 0.992 |
| Wavelet_HL_GLDM_DependenceNonUniformity | 14 | 5 | 0.196 | 1 |
| Wavelet_HL_GLRLM_LongRunEmphasis | 14 | 2 | -0.200 | 1 |
| Wavelet_HL_GLRLM_RunVariance | 14 | 2 | -0.163 | 0.992 |
| Wavelet_LH_FirstOrder_10Percentile | 14 | 4 | -0.187 | 1 |
| Wavelet_LL_GLSZM_GrayLevelNonUniformity | 14 | 2 | -0.211 | 1 |
| Wavelet_LL_GLSZM_LargeAreaHighGrayLevelEmphasis | 14 | 3 | -0.154 | 0.992 |
| LoG-3mm_GLCM_Idmn | 15 | 2 | 0.162 | 0.784 |
| LoG-5mm_FirstOrder_Kurtosis | 15 | 3 | 0.183 | 0.768 |
| LoG-5mm_GLCM_Idmn | 15 | 2 | 0.256 | 0.88 |
| LoG-5mm_GLRLM_LongRunHighGrayLevelEmphasis | 15 | 2 | -0.172 | 0.808 |
| LoG-5mm_GLSZM_LargeAreaEmphasis | 15 | 5 | -0.171 | 0.704 |
| LoG-5mm_GLSZM_LargeAreaLowGrayLevelEmphasis | 15 | 3 | 0.212 | 0.8 |
| LoG-5mm_GLSZM_ZoneVariance | 15 | 5 | -0.159 | 0.704 |
| Original_GLRLM_LongRunLowGrayLevelEmphasis | 15 | 4 | 0.160 | 0.752 |
| Wavelet_LL_GLRLM_LongRunLowGrayLevelEmphasis | 15 | 4 | 0.196 | 0.784 |
| Wavelet_LL_GLSZM_LargeAreaHighGrayLevelEmphasis | 15 | 3 | 0.206 | 0.8 |
| LoG-3mm_GLDM_GrayLevelNonUniformity | 16 | 3 | 0.163 | 0.328 |
| LoG-3mm_GLRLM_ShortRunHighGrayLevelEmphasis | 16 | 2 | 0.166 | 0.624 |
| LoG-5mm_GLRLM_RunEntropy | 16 | 4 | 0.307 | 0.68 |
| Wavelet_HH_GLRLM_RunLengthNonUniformity | 16 | 5 | -0.285 | 0.76 |
| Wavelet_HL_FirstOrder_Kurtosis | 16 | 2 | 0.371 | 0.36 |
| Wavelet_HL_GLSZM_LargeAreaHighGrayLevelEmphasis | 16 | 3 | 0.237 | 0.352 |
| Wavelet_LL_GLCM_InverseVariance | 16 | 3 | -0.223 | 0.2 |
| Wavelet_LL_GLDM_DependenceNonUniformity | 16 | 4 | 0.195 | 0.312 |
| Wavelet_LL_GLDM_DependenceNonUniformityNormalized | 16 | 2 | -0.186 | 0.28 |
| Wavelet_LL_GLDM_DependenceVariance | 16 | 5 | 0.304 | 0.312 |
| LoG-3mm_GLDM_SmallDependenceHighGrayLevelEmphasis | 17 | 5 | 0.151 | 0.992 |
| LoG-3mm_GLRLM_ShortRunHighGrayLevelEmphasis | 17 | 2 | 0.161 | 0.992 |

| | | | | |
|---|---|---|---|---|
| LoG-3mm_GLSZM_ZonePercentage | 17 | 5 | 0.151 | 1 |
| Wavelet_HH_GLSZM_LargeAreaLowGrayLevelEmphasis | 17 | 5 | 0.153 | 1 |
| Wavelet_HH_GLSZM_ZoneVariance | 17 | 5 | 0.154 | 0.976 |
| Wavelet_HL_FirstOrder_Range | 17 | 3 | -0.159 | 1 |
| Wavelet_HL_GLDM_DependenceEntropy | 17 | 5 | -0.199 | 0.984 |
| Wavelet_HL_GLDM_DependenceVariance | 17 | 5 | -0.194 | 0.976 |
| Wavelet_HL_GLRLM_GrayLevelNonUniformity | 17 | 2 | -0.212 | 1 |
| LoG-5mm_GLDM_LargeDependenceLowGrayLevelEmphasis | 18 | 2 | -0.106 | 0.016 |
| LoG-5mm_GLRLM_LongRunHighGrayLevelEmphasis | 18 | 2 | -0.994 | 0.312 |
| LoG-1mm_NGTDM_Strength | 19 | 5 | -0.229 | 0.824 |
| LoG-3mm_GLRLM_RunEntropy | 19 | 5 | 0.268 | 0.72 |
| LoG-3mm_GLSZM_LargeAreaHighGrayLevelEmphasis | 19 | 5 | -0.165 | 0.808 |
| Original_FirstOrder_10Percentile | 19 | 2 | 0.194 | 0.792 |
| Original_GLDM_DependenceNonUniformity | 19 | 2 | 0.296 | 0.816 |
| Original_GLRLM_LongRunLowGrayLevelEmphasis | 19 | 4 | -0.170 | 0.776 |
| Original_GLSZM_LargeAreaEmphasis | 19 | 4 | -0.178 | 0.728 |
| Wavelet_HL_GLSZM_ZonePercentage | 19 | 5 | -0.209 | 0.808 |
| Wavelet_LH_NGTDM_Busyness | 19 | 3 | 0.158 | 0.72 |
| Wavelet_LL_GLCM_InverseVariance | 19 | 3 | 0.160 | 0.768 |
| LoG-1mm_GLRLM_GrayLevelNonUniformity | 20 | 2 | 0.154 | 0.992 |
| LoG-3mm_FirstOrder_90Percentile | 20 | 5 | 0.154 | 0.944 |
| LoG-3mm_FirstOrder_Skewness | 20 | 3 | 0.166 | 0.984 |
| LoG-5mm_GLRLM_LongRunLowGrayLevelEmphasis | 20 | 2 | 0.146 | 0.96 |
| Original_GLDM_DependenceNonUniformity | 20 | 2 | 0.152 | 0.96 |
| Original_Sphericity | 20 | 2 | 0.192 | 0.968 |
| Wavelet_HL_FirstOrder_Kurtosis | 20 | 2 | -0.211 | 0.968 |
| Wavelet_LH_FirstOrder_Maximum | 20 | 5 | 0.148 | 0.952 |
| Wavelet_LH_GLDM_GrayLevelNonUniformity | 20 | 2 | 0.185 | 0.976 |
| Wavelet_LH_GLRLM_RunVariance | 20 | 2 | -0.156 | 0.952 |
| LoG-1mm_GLSZM_LargeAreaHighGrayLevelEmphasis | 21 | 2 | 0.151 | 0.736 |
| LoG-3mm_FirstOrder_Skewness | 21 | 3 | 0.208 | 0.856 |
| LoG-5mm_GLCM_ClusterShade | 21 | 2 | -0.158 | 0.848 |
| LoG-5mm_GLSZM_SizeZoneNonUniformity | 21 | 5 | -0.236 | 0.864 |
| Original_GLSZM_SizeZoneNonUniformityNormalized | 21 | 5 | -0.244 | 0.88 |
| Wavelet_HH_GLSZM_LargeAreaHighGrayLevelEmphasis | 21 | 5 | 0.255 | 0.84 |
| Wavelet_HH_GLSZM_LargeAreaLowGrayLevelEmphasis | 21 | 5 | 0.262 | 0.856 |
| Wavelet_HH_GLSZM_ZoneVariance | 21 | 5 | 0.271 | 0.824 |
| Wavelet_HL_NGTDM_Complexity | 21 | 3 | 0.158 | 0.672 |
| Wavelet_LH_GLRLM_LongRunLowGrayLevelEmphasis | 21 | 2 | 0.198 | 0.856 |
| LoG-3mm_GLSZM_LargeAreaHighGrayLevelEmphasis | 22 | 5 | -0.161 | 0.696 |
| LoG-5mm_FirstOrder_TotalEnergy | 22 | 2 | 0.152 | 0.728 |
| LoG-5mm_GLSZM_LargeAreaEmphasis | 22 | 5 | 0.168 | 0.608 |
| LoG-5mm_GLSZM_LargeAreaLowGrayLevelEmphasis | 22 | 3 | -0.169 | 0.696 |

| | | | | |
|---|---|---|---|---|
| LoG-5mm_GLSZM_SizeZoneNonUniformity | 22 | 5 | -0.165 | 0.832 |
| LoG-5mm_GLSZM_ZoneEntropy | 22 | 5 | 0.171 | 0.712 |
| LoG-5mm_GLSZM_ZoneVariance | 22 | 5 | 0.161 | 0.632 |
| Wavelet_HH_GLSZM_LargeAreaHighGrayLevelEmphasis | 22 | 5 | -0.416 | 0.72 |
| Wavelet_HH_GLSZM_LargeAreaLowGrayLevelEmphasis | 22 | 5 | -0.362 | 0.76 |
| Wavelet_HH_GLSZM_ZoneVariance | 22 | 5 | -0.408 | 0.72 |
| LoG-1mm_GLSZM_LargeAreaHighGrayLevelEmphasis | 23 | 2 | -0.176 | 1 |
| LoG-3mm_GLSZM_LargeAreaHighGrayLevelEmphasis | 23 | 5 | -0.181 | 0.976 |
| LoG-5mm_GLSZM_SizeZoneNonUniformity | 23 | 5 | -0.149 | 0.984 |
| LoG-5mm_NGTDM_Contrast | 23 | 2 | 0.145 | 0.992 |
| Original_GLDM_LargeDependenceLowGrayLevelEmphasis | 23 | 5 | -0.206 | 0.992 |
| Original_GLDM_LowGrayLevelEmphasis | 23 | 5 | -0.217 | 0.992 |
| Original_GLDM_SmallDependenceLowGrayLevelEmphasis | 23 | 5 | -0.232 | 1 |
| Original_GLSZM_SizeZoneNonUniformityNormalized | 23 | 5 | -0.147 | 0.984 |
| Wavelet_LL_GLDM_DependenceVariance | 23 | 5 | -0.148 | 0.984 |
| Wavelet_LL_GLDM_SmallDependenceLowGrayLevelEmphasis | 23 | 5 | -0.150 | 1 |
